# Long-term Exposures to Air Pollutants Affect FeNO in Children: A Longitudinal Study

**DOI:** 10.1101/2021.03.01.21252712

**Authors:** Yue Zhang, Sandrah P. Eckel, Kiros Berhane, Erika Garcia, Patrick Muchmore, Noa Ben-Ari Molshatzki, Edward B. Rappaport, William S. Linn, Rima Habre, Frank D. Gilliland

## Abstract

Fractional exhaled nitric oxide (FeNO) is a marker of airway inflammation shown to be responsive to short-term air pollution exposures; however, effects of long-term exposures are uncertain. Using longitudinal assessments of FeNO and air pollutant exposures, we aimed to determine whether FeNO is a marker for chronic effects of air pollution exposures after accounting for short-term exposures effects.

FeNO was assessed up to six times 2004-2012 in 3607 schoolchildren from 12 communities in the Southern California Children’s Health Study. Within-community long-term ambient air pollution exposures (PM_2.5_, PM_10_, NO_2_, O_3_) were represented by differences between community-specific annual averages and the eight-year average spanning the study period. Linear mixed-effect models estimated within-participant associations of annual average air pollution with current FeNO, controlling for previous FeNO, prior seven-day average pollution, potential confounders, and community-level random intercepts. We considered effect modification by sex, ethnicity, asthma, and allergy at baseline.

We found FeNO was positively associated with annual average air pollution, after accounting for short-term exposures. One standard deviation higher annual PM_2.5_ and NO_2_ exposures (PM_2.5:_2.0 μg/m^3^ ; NO_2_:2.7 ppb) were associated, respectively, with 4.6% (95%CI:2.3%-6.8%) and 6.5% (95%CI:4.1%-8.9%) higher FeNO. These associations were larger among females. We found little evidence supporting association with PM_10_ or O_3_.

Annual average PM_2.5_ and NO_2_ levels were associated with FeNO in schoolchildren, adding new evidence that long-term exposure affects FeNO beyond the well-documented short-term effects. Longitudinal FeNO measurements may be useful as an early marker of chronic respiratory effects of long-term PM_2.5_ and NO_2_ exposures in children.

**Key messages:**

- We show strong evidence that long-term exposures to air pollutants affect FeNO, independent of the well-documented associations with short-term exposures to air pollution
- Longitudinal FeNO measurements may be useful as an early marker of chronic respiratory effects of long-term air pollution exposures in children.

**Capsule summary:** Annual average PM_2.5_ and NO_2_ were associated with FeNO in schoolchildren, adding new evidence that long-term exposure affects FeNO beyond the well-documented short-term effects.

## INTRODUCTION

Breathing polluted air has a multitude of impacts on human health, many of which are thought to be driven by inflammatory mechanisms. The primary route of exposure is inhalation, thus the respiratory system is a major target organ for toxic effects as it receives the largest dose. The fractional concentration of exhaled nitric oxide (FeNO) is a marker of airway inflammation which has been shown to be useful for the assessment of acute respiratory effects of short-term exposure (hours to weeks) to ambient air pollutants,^1-3^ including particulate matter with an aerodynamic diameter of 2.5 or 10 μm or less (PM_2.5_ and PM_10_, respectively), black smoke, elemental carbon (EC), nitrogen dioxide (NO_2_), and ultrafine particles. In the Southern California Children’s Health study (CHS), we have reported positive cross-sectional associations of short-term average PM_10_, PM_2.5_, and ozone (O_3_) exposures in the week(s) before FeNO measurement with FeNO.^4^

Currently, there is limited and inconsistent evidence for effects of longer-term air pollution exposures on FeNO. For example, a number of cross-sectional studies have related FeNO to either location-based proxies for long-term exposures to traffic/industrial activity or long-term (seasonal to annual average) ambient pollution. Some of these studies found significant positive associations^5-7^ (two with adjustment for short-term exposures^8, 9^) but others did not.^10-15^ An early analysis of CHS data over a short follow-up period is one of the few studies to date relating longitudinally assessed FeNO to long-term ambient air pollution exposures. Using two assessments of FeNO approximately one year apart in ∼1200 schoolchildren, we found that increased annual average PM_2.5_ or NO_2_ was associated with increased within-participant FeNO, after adjusting for short-term exposures (in the prior week).^16^

We hypothesize that FeNO is affected by both long-term and short-term exposures to ambient air pollution. In this study, the estimand of interest is long-term ambient air pollution exposure effect, and short-term exposure is treated as nuisance parameter. We examined the full longitudinal CHS data—which includes up to six repeated FeNO assessments in more than 3000 schoolchildren—to investigate whether long-term (i.e., annual average) ambient air pollution exposures are associated with within-participant changes in FeNO, after accounting for short-term (i.e., weekly average) exposures.

## MATERIAL AND METHODS

### Study Population

Participants in this study were part of the most recent southern California Children’s Health Study (CHS) cohort, which started enrollment in 2002-2003 from kindergarten and first grade classrooms. Detailed information on study design, participant recruitment, and data collection has been reported previously.^17^ This analysis includes 3607 school children aged 6-17 years from 12 communities in Southern California, who had FeNO assessed up to 6 times in approximately annual visits from 2004-2005 through 2011-2012. Ethical approval of the protocol was obtained from the University of Southern California institutional review board (approval # HS-13-00150, continuing review date April 17, 2019). Informed assent was obtained for each child, and informed consent from a parent or guardian. The research protocol was reviewed and approved by the University of Southern California Health Campus Institutional Review Board.

### FeNO measurement

FeNO was measured at schools following the American Thoracic Society (ATS) guidelines using an offline technique at the first two visits (2004-2005 and 2005-2006 school years) and online technique in the subsequent four visits. For offline measurement, breath was collected in aluminized Mylar bags using commercial breath sampling kits (Sievers Division, GE Analytical Instruments, Boulder, Colorado, USA). Each child took 2-3 initial deep breaths through the kit’s NO scrubber followed by a near-vital-capacity breath and immediate controlled exhalation through the sampling kit. The collected bag samples were measured in central laboratory for FeNO level at targeted 100 ml/sec flow rate using a standard chemiluminescent analyzer system (Sievers Division). For online measurement, FeNO was measured directly using EcoMedics CLD-88-SP analyzers with DeNOx accessories to provide NO-free inhaled air (EcoPhysics Inc, Ann Arbor, MI, USA/Duernten, Switzerland). Each child was asked to provide at least three maneuvers at the target 50 ml/sec flow rate. The plateau concentration for each maneuver was taken from the 3 second plateau with the minimum NO coefficient of variation. FeNO was represented by the mean of two or three ATS acceptable plateaus that differed by less than 15% or 1 *ppb* where all ATS acceptable plateaus were less than 10 *ppb*. Offline FeNO data were converted to estimates of FeNO as would be measured online at 50 ml/sec expiratory flow. The conversion was performed using a statistical model determined in the post hoc comparison study, which involved 362 children measured on and off line in the same testing session. We exclude observations with reported use of inhaled corticosteroids. Details of FeNO data collection, quality control, and conversion procedures have been reported previously.^18-20^

### Air Pollution Data

Daily 24-hour average PM_2.5_, PM_10_, NO_2_, O_3_ (10am-6pm only), and temperature data were obtained for the study period from central monitoring sites in each community operated by local air pollution agencies in conformance with US Environmental Protection Agency (EPA) requirements. For each pollutant, community-specific study period averages were computed (a seven-year average, from 2004/05-2011/12). For each pollutant and each participants’ FeNO test date, annual average pollution was calculated based on the 12 months preceding the test date in that participant’s community. Weekly averages were calculated based on the 7 days preceding the test date. Air pollution data was interpolated to fill in data gaps on days with missing exposure using modeled predictions based on data from nearby monitors. While such processes could be potential sources of bias, previous sensitivity analyses on CHS data that limited use to complete data have not altered main study findings.^4^ The primary “long-term” exposure of interest for this study was the annual within-community fluctuation, defined as the difference between annual averages and the seven year average concentration for each community. We refer to seven day average as “short-term” exposures hereafter.

### Other Covariates

Race/ethnicity, medication use, current wheeze status, exposure to secondhand tobacco smoke (SHS), asthma history, and allergy history were determined from annual self-administered questionnaires. Questionnaires were completed by parents from baseline to year 2006-2007, and by children from follow-up year 2007-2008 onwards. Questionnaires were collected at the school visit during which the child’s FeNO test was performed. Ambient air nitric oxide was measured at the time of FeNO collection. Trained technicians measured height and weight at every FeNO test date following a standardized protocol. Age- and sex-specific body mass index (BMI) percentiles were calculated based on the Centers for Disease Control and Prevention BMI growth charts^21^.

### Statistical Analysis

FeNO had a right-skewed distribution, so we used geometric mean and geometric standard deviation to quantify central tendency and dispersion and performed all analyses with natural log-transformed FeNO. In descriptive data analyses, we summarized the characteristics of the study participants, compared the distribution of baseline year log FeNO by these characteristics, and also summarized the distribution of the community-specific seven-year pollutant averages and the annual within-community fluctuations. We also calculated Pearson correlation coefficients of annual within-community fluctuations between pairs of pollutants.

We estimated linear mixed effect models to examine the association of log FeNO with annual within-community fluctuations in air pollution, adjusting for short-term exposures, confounders, and design variables as well as log FeNO at the previous visit. More details on the model specification could be found in the Supplementary appendix. Potential confounders included sex, race/ethnicity, and respiratory allergy at baseline, as well as time-varying measures of asthma status, medication use, current wheeze, secondhand tobacco smoke exposure, recent respiratory illness, concurrent room air nitric oxide, month of FeNO test, and average temperature in the week preceding the FeNO test. To assess confounding, we used the analytic change-in-estimate (2:’10%) criterion. We also evaluated whether long-term exposure associations were modified by: baseline asthma, baseline allergy, sex, and ethnicity. In sensitivity analyses, the final models were refitted by: (a) removing the five communities which contributed to only the first four (of six) FeNO visits, (b) removing one community at a time, (c) adding a community-level random slope on long-term exposure to investigate potential heterogeneity by community, (d) subsetting to only online FeNO collection (the last four of the six visits), or (e) accounting for the serial correlation of FeNO observations from same participant by using an autoregressive within-participant level random component instead of controlling for previous visit log FeNO. In all models, missing data were assumed to be missing at random. All analyses were conducted using the statistical package R version 3.5.1^22^. Hypothesis tests were performed under a 0.05 significance level and a two-sided alternative.

## RESULTS

The 3607 children, with mean age 9 at baseline, were evenly distributed by sex (male: 49.7%), primarily Hispanic (56.3%) or non-Hispanic White (33.2%), 41.5% had allergies, 15.1% had asthma, and 3% had reported a recent respiratory illness (Table 1). At baseline, the geometric mean FeNO was 12.1 ppb (geometric SD: 1.9 ppb) and baseline FeNO was statistically significantly associated with most participant characteristics. Specifically, boys on average had significantly higher FeNO than girls (p-value=0.015). FeNO differed by race/ethnicity (p-value <0.001), with African American children having higher FeNO than the other race/ethnicity groups. Children with respiratory conditions, such as asthma, wheeze, allergy, and recent respiratory illness, had statistically significantly higher FeNO than those without (p-values all <0.001). FeNO was positively associated with age and height (p-value <0.001), but not BMI (p-value=0.831). FeNO varied by month of collection (p-value<0.001), with the lowest geometric means in cooler months (December – March).

**Table 1:**
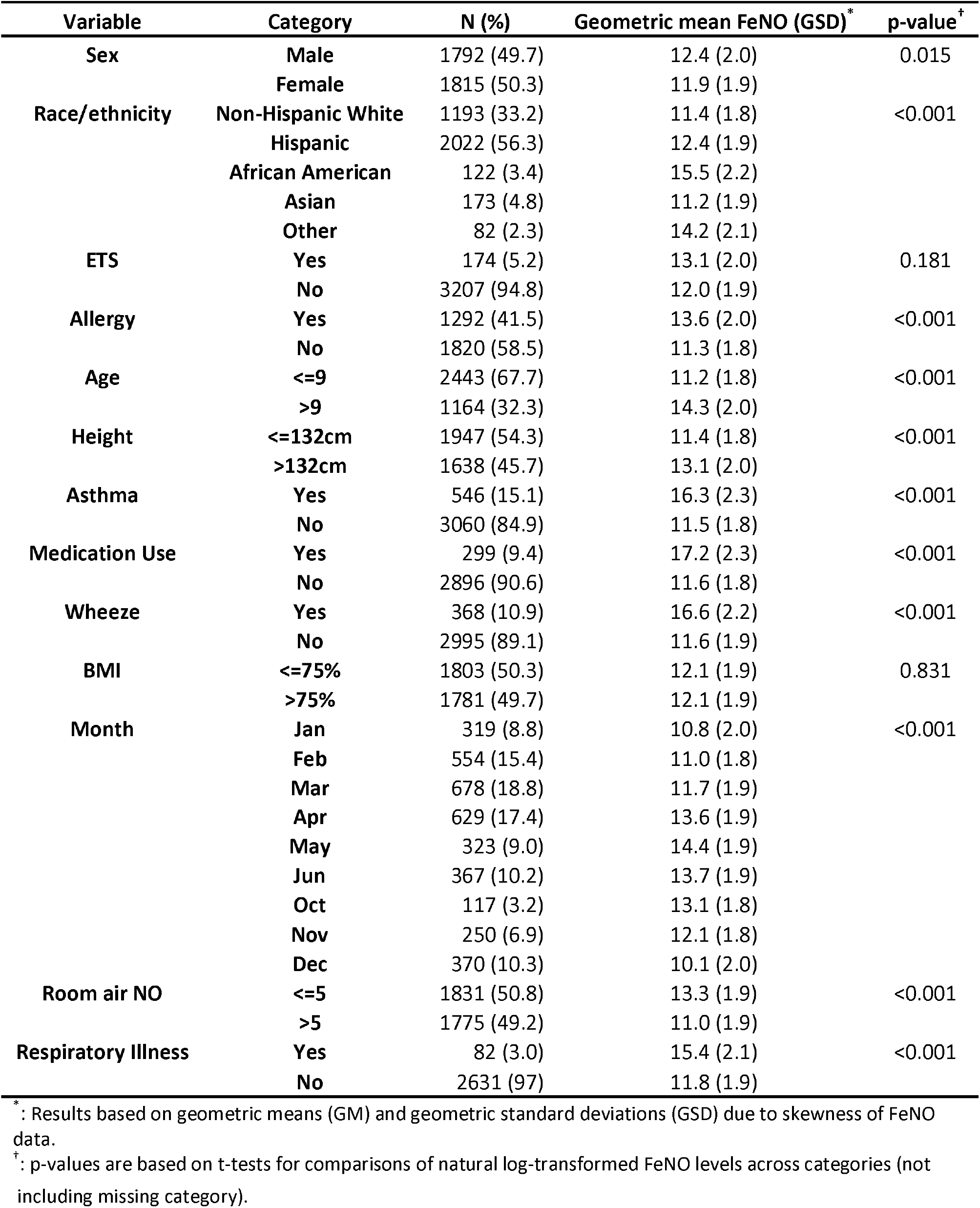
Summary of Participant Characteristics and FeNO at Baseline (N=3607)

Community-specific seven-year average air pollution concentrations ranged approximately two to three-fold across the 12 communities (Table 2). Most communities experienced a downward trend in annual average PM_2.5_ and NO_2_ over the study period (Figure 1), which was a major source of variation in the annual within-community fluctuations for these pollutants. The standard deviation of annual within-community fluctuations was 2.0 µg/m^3^ for PM_2.5_, 6.7 µg/m^3^ for PM_10_, 2.7 ppb for NO_2_, and 2.5 ppb for O_3_ (Table 2). As shown in Table 3, PM_2.5_ and PM_10_ had the highest correlation in seven-year averages across-communities (Pearson’s R=0.81), while PM_2.5_ and NO_2_ had the largest correlation in annual within-community fluctuations (Pearson’s R=0.64) and annual within-community fluctuations of O_3_ were negatively correlated with the other pollutants. Log FeNO was correlated with log FeNO at the previous visit (Pearson’s R=0.65), indicating moderately strong first order autocorrelation.

**Figure 1:**
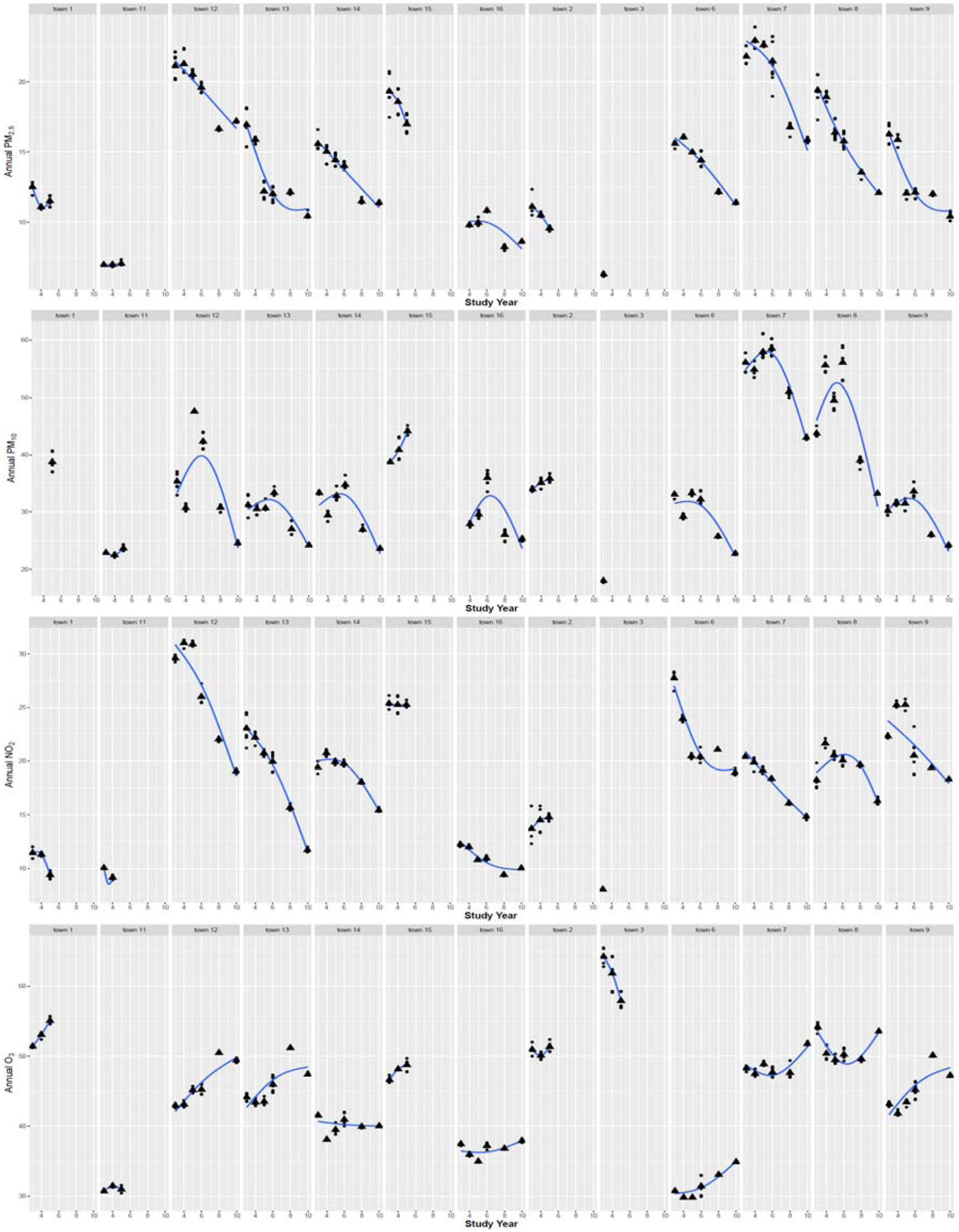
Community-specific annual average air pollution during the study period. Note: 1=Alpine; 2=Lake Elsinore; 3=Lake Gregory; 6=Long Beach; 7=Mira Loma; 8=Riverside; 9=San Dimas; 11=Santa Maria; 12=Upland; 13=Glendora; 14=Anaheim; 15=San Bernardino; 16=Santa Barbara. Study Year 3=2004-2005; Study Year 4=2005-2006; Study Year 5=2006-2007; Study Year 6=2007-2008; Study Year 7=2008-2009; Study Year 8=2009-2010; Study Year 9=2010-2011; Study Year 10=2011-2012; The dot points are the annual average air pollution level prior to the dates of FeNO measurement, and triangle points are the annual average air pollution level at each study year. The curve lines are the fitted trends of annual average air pollution level across the study years for each community.

**Table 2:** Distribution of the 7-year Average Air Pollutant Concentrations and Annual Within-Community Fluctuation across 12 Communities.

| Pollutant<sup>#</sup> | 7-year Average |  | Annual Within-Community Fluctuation |  |
| --- | --- | --- | --- | --- |
|  | Mean (SD) | Min-Max | Mean (SD) | Min-Max |
| PM<sub>2.5</sub>, µg/m<sup>3</sup> | 12.9 (3.8) | 6.8-19.4 | 0.1 (2.0) | -3.8-4.8 |
| PM<sub>10</sub>, µg/m<sup>3</sup> | 32.0 (8.3) | 22.5-52.7 | 2.1 (6.7) | -10.2-23.3 |
| NO<sub>2</sub>, ppb | 16.7 (6.3) | 6.7-25.6 | -0.1 (2.7) | -7.2-5.7 |
| O<sub>3</sub>, ppb | 45.1 (8.4) | 30.7-56.6 | -0.1 (2.5) | -6.1-7.7 |

<sup>#</sup>: All pollutants are 24 hr average, except for ozone (10am–6pm average).

**Table 3:**
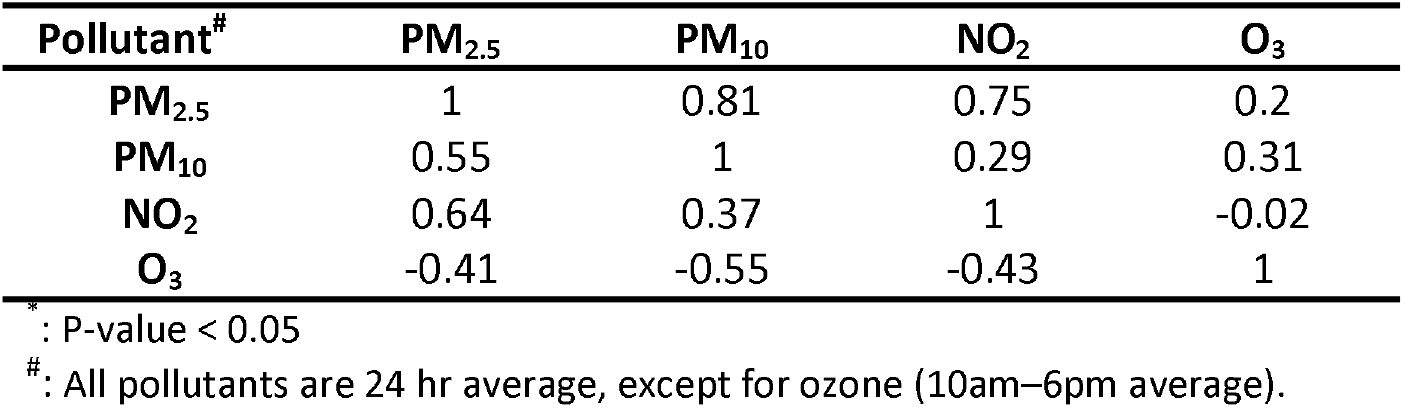
Pearson Correlation Coefficients of Annual Within-Community Fluctuation in Air Pollution (lower triangular matrix) and 7-year average air pollutant concentrations (upper triangular matrix).

| Pollutant<sup>#</sup> | PM<sub>2.5</sub> | PM<sub>10</sub> | NO<sub>2</sub> | O<sub>3</sub> |
| --- | --- | --- | --- | --- |
| PM<sub>2.5</sub> | 1 | 0.81 | 0.75 | 0.2 |
| PM<sub>10</sub> | 0.55 | 1 | 0.29 | 0.31 |
| NO<sub>2</sub> | 0.64 | 0.37 | 1 | -0.02 |
| O<sub>3</sub> | -0.41 | -0.55 | -0.43 | 1 |

<sup>\*</sup>: P-value < 0.05
<sup>#</sup>: All pollutants are 24 hr average, except for ozone (10am–6pm average).

Long-term exposures to PM_2.5_ and NO_2_ were positively associated with within-person changes in FeNO, after adjustment for covariates and short-term exposure (Table 4). A one SD (2.0 μg/m^3^) increase in the annual within-community fluctuation of PM_2.5_ was associated with a 4.6% within-participant increase in FeNO (95% CI: 2.3%-6.8%), controlling for covariates including FeNO at the previous visit and seven-day average PM_2.5_. Similarly, a one SD (2.7 ppb) increase in the annual within-community fluctuation of NO_2_ was associated with a 6.5% within-participant increase in FeNO (95% CI: 4.1%-8.9%), controlling for covariates. We found no evidence for an association of long-term PM_10_ and FeNO. Long-term O_3_ was negatively associated with FeNO in the primary analysis, but this association was attenuated and no longer statistically significant in sensitivity analyses (Tables 1S and 2S). All models were adjusted for sex, race, respiratory allergy, asthma, medication use, wheeze, secondhand tobacco smoking, recent respiratory illness, room air nitric oxide, month, temperature, and short-term exposure of air pollution.

**Table 4:**
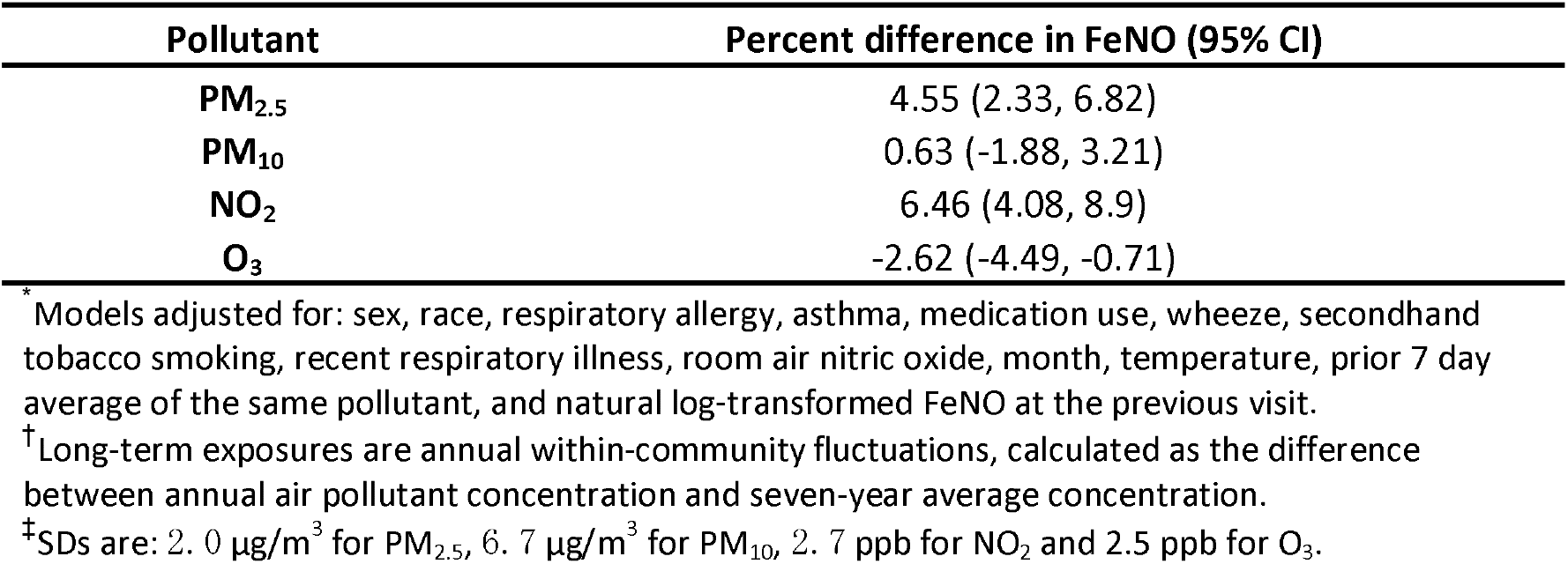
Adjusted^*^ percent difference in FeNO associated with a one standard deviation (SD)^‡^ increase in long-term air pollution exposures^†^.

There was evidence that the associations of long-term PM_2.5_ and NO_2_ with FeNO varied by sex, with larger estimated associations in females (interaction p-values: 0.007 for PM_2.5_ and 0.066 for NO_2_) (Table 5). In sensitivity analyses (Supplementary Table 1), we still observed statistically significant positive associations of long-term PM_2.5_ and NO_2_ with FeNO after: (a) removing the five communities followed only through four years, (b) removing one community at a time (Supplementary Table 2), and (c) taking out the adjustment for short-term air pollution, though we did observe some heterogeneity in the associations.

**Table 5:**
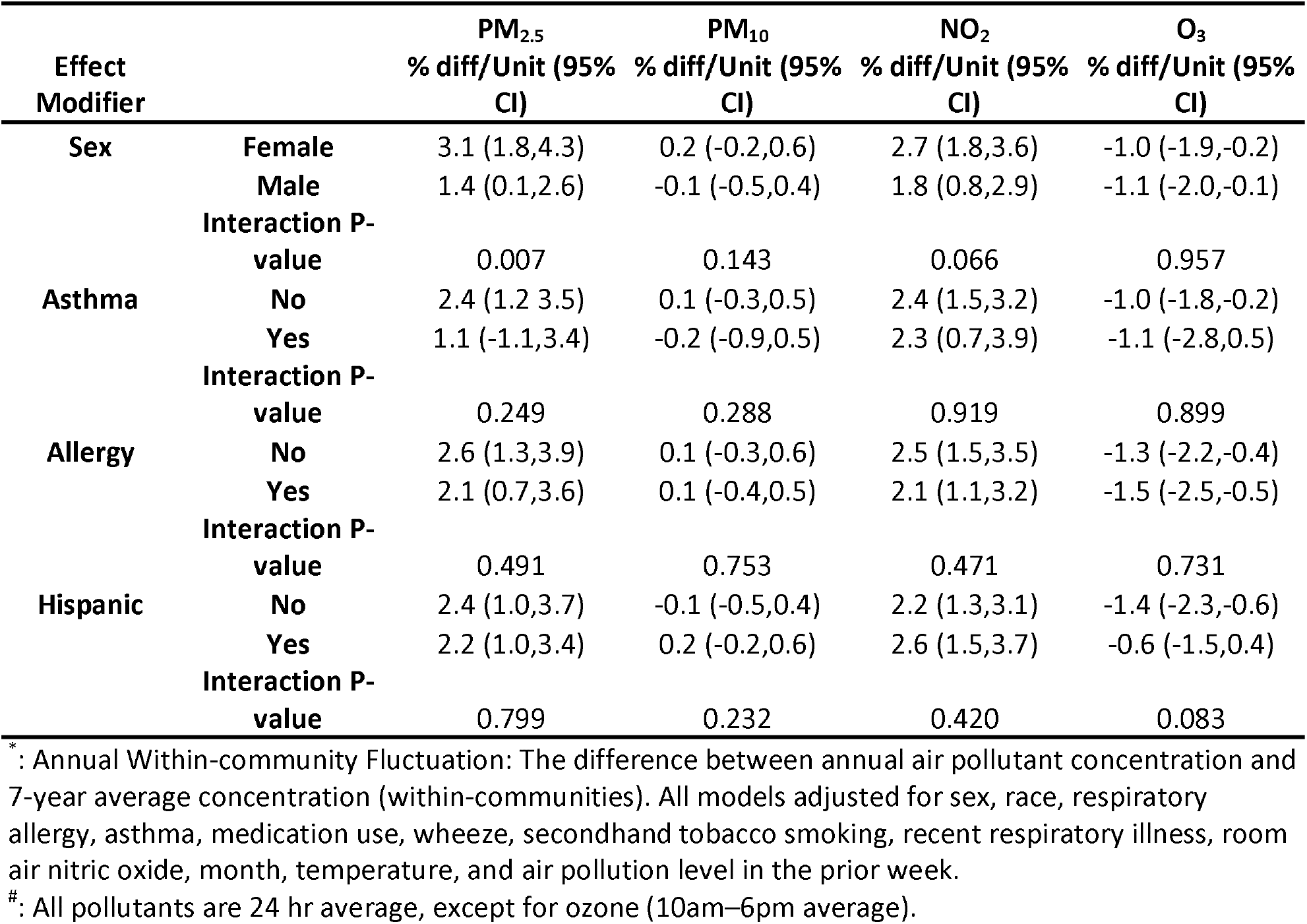
Interaction Tests^*^ on the Effects of Annual Within-Community Fluctuation in Air Pollutions^#^ on FeNO level.

| Effect Modifier |  | PM<sub>2.5</sub><br>% diff/Unit (95% CI) | PM<sub>10</sub><br>% diff/Unit (95% CI) | NO<sub>2</sub><br>% diff/Unit (95% CI) | O<sub>3</sub><br>% diff/Unit (95% CI) |
| --- | --- | --- | --- | --- | --- |
| Sex | Female | 3.1 (1.8,4.3) | 0.2 (-0.2,0.6) | 2.7 (1.8,3.6) | -1.0 (-1.9,-0.2) |
|  | Male | 1.4 (0.1,2.6) | -0.1 (-0.5,0.4) | 1.8 (0.8,2.9) | -1.1 (-2.0,-0.1) |
| Interaction P-value |  | 0.007 | 0.143 | 0.066 | 0.957 |
| Asthma | No | 2.4 (1.2,3.5) | 0.1 (-0.3,0.5) | 2.4 (1.5,3.2) | -1.0 (-1.8,-0.2) |
|  | Yes | 1.1 (-1.1,3.4) | -0.2 (-0.9,0.5) | 2.3 (0.7,3.9) | -1.1 (-2.8,0.5) |
| Interaction P-value |  | 0.249 | 0.288 | 0.919 | 0.899 |
| Allergy | No | 2.6 (1.3,3.9) | 0.1 (-0.3,0.6) | 2.5 (1.5,3.5) | -1.3 (-2.2,-0.4) |
|  | Yes | 2.1 (0.7,3.6) | 0.1 (-0.4,0.5) | 2.1 (1.1,3.2) | -1.5 (-2.5,-0.5) |
| Interaction P-value |  | 0.491 | 0.753 | 0.471 | 0.731 |
| Hispanic | No | 2.4 (1.0,3.7) | -0.1 (-0.5,0.4) | 2.2 (1.3,3.1) | -1.4 (-2.3,-0.6) |
|  | Yes | 2.2 (1.0,3.4) | 0.2 (-0.2,0.6) | 2.6 (1.5,3.7) | -0.6 (-1.5,0.4) |
| Interaction P-value |  | 0.799 | 0.232 | 0.420 | 0.083 |

\*: Annual Within-community Fluctuation: The difference between annual air pollutant concentration and 7-year average concentration (within-communities). All models adjusted for sex, race, respiratory allergy, asthma, medication use, wheeze, secondhand tobacco smoking, recent respiratory illness, room air nitric oxide, month, temperature, and air pollution level in the prior week.
#: All pollutants are 24 hr average, except for ozone (10am–6pm average).

## DISCUSSION

In a large, prospective cohort study of schoolchildren in 12 Southern California communities, we found statistically significant positive associations of FeNO—assessed longitudinally over an eight-year period—with within-community fluctuations in annual average PM_2.5_ and NO_2_ after adjusting for prior week exposure, previous visit FeNO, and confounders. This provides the strong evidence that long-term exposures to air pollutants affect FeNO, independent of the well-documented associations with short-term exposures to air pollution.

Results of this longitudinal study (using all six FeNO visits in the CHS) are consistent with a preliminary longitudinal analysis of CHS data (using only the first two online FeNO assessments),^16^ which had found *within-participant* associations of long-term PM_2.5_ and NO_2_ (but not PM_10_ or O_3_) with FeNO while adjusting for short-term exposure. The preliminary CHS study had targeted within-participant changes in FeNO using a different but complementary statistical approach in which the change in FeNO was modeled as a function of the change in annual average air pollution.^16^ When studying FeNO, it is important to consider within-participant changes over time from longitudinally collected data, given the considerable unexplained across-participant heterogeneity in FeNO typically observed in cross-sectional studies (e.g., R^2^<0.3).^23^ Indeed, in our data we observed moderately strong autocorrelation in FeNO indicating relatively stable within-person FeNO. As an additional example, in 158 asthmatic adults, the intraclass correlation of FeNO over one year was 0.73, with low baseline FeNO tending to stay low and higher baseline FeNO tending to stay high, but with more variability.^24^ To our knowledge, the only other study relating longitudinally assessed FeNO to long-term air pollution exposures found positive but non-statistically-significant associations of FeNO (measured twice over six months in 130 children) with seasonal black carbon levels at subjects’ residence (estimated by land-use regression), even after adjusting for short-term exposures (2 hour, 24 hour, or 7 day average black carbon from central site monitors).^25^ This study’s statistical analysis did not fully target within-participant associations since it used linear mixed models with a participant-level random intercept but did not decompose time-varying exposures (X_ij_) into within- and between-person terms (i.e., 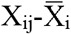. and 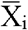.).^26^

A number of cross-sectional studies have related FeNO to long-term (seasonal to annual average) ambient pollution or location-based proxies for long-term exposures to traffic/industrial activity. By design, these studies can estimate only between-participant associations of exposures with FeNO and hence are subject to the limitations of this approach. Some cross-sectional studies provide evidence supporting long-term exposure effects. In 241 healthy, full-term infants, prenatal average NO_2_ from a central monitoring site was associated with increased FeNO measured during sleep at five weeks of age, even after adjusting for mean NO_2_ in the 2 days prior to the FeNO test or residential distance to major road.^9^ Two studies relating FeNO to proximity to industrial activity found FeNO to be elevated in children living closer to oil refinery/petrochemical activity^6^ and in children attending schools closer to a steel mill,^7^ though neither study adjusted for short-term air pollution exposures. Three studies^5, 8, 27^ found a similar magnitude positive association between a proxy measure for traffic exposure (length of road in a circular buffer around the home), with the strongest associations for the smallest buffers around the home. While the length of road traffic exposure proxy likely represents both long- and short-term exposures, one of these studies adjusted for short-term exposure (central site 24 hour average ambient pollution) and found the length of road association to be robust to the adjustment,^8^ suggesting a long-term exposure effect. Other cross-sectional studies have found no evidence for long-term exposure effects, perhaps due to exposure metrics relying on spatial variation which are not sensitive to traffic sources on smaller, local roads (e.g., land-use regression predicted seasonal or annual averages,^14, 15, 25^ traffic counts,^10, 13^ or distance to major road^11, 13^) or too few locations with high/low exposure to separate the source of interest from potential confounders.^12^

Many study communities observed declines in annual average PM_2.5_ and NO_2_ during the study period. These declines were a key source of the variation in our long-term exposure metric (within-community annual fluctuations, calculated as the difference between annual averages and the seven-year average concentration for each community). Our standard interpretation of the observed positive associations as “percent *increase* in FeNO per SD *increase* in exposure” (e.g., 6.5% increase in FeNO (95% CI: 4.1%-8.9%) per 2.7 ppb increase in long-term NO_2_), could alternatively be reformulated as a “percent *decrease* in FeNO per SD *decrease* in exposure”. For example, scaled to an observed declines, in Glendora annual average NO_2_ decreased by 11.3 ppb over the study period, and we estimated a 30% *decrease* in FeNO (95% CI: 18.2% to 42.9%) per 11.3 ppb *decrease* in long-term NO_2_). A number of studies have used natural or designed experiments to demonstrate that FeNO decreased following decreasing (primarily short-term) exposures. For example, following the adoption of clean fuel/technology school buses, 10-30% reductions in FeNO were observed in 275 schoolchildren.^28^ Followingdramatic drops in air pollution in response to anti-air pollution measures enacted for the Beijing Olympics, FeNO decreased ∼57% in 125 health young adults (and subsequently rose 135% after the games)^3^ and ∼27% in 36 children.^29^ Within-participant FeNO also declined in a group of 42 retired teachers relocated from a higher air pollution city to a lower pollution city for 9 days^30^ and in 37 untreated allergic children moved from an urban to a rural environment for 7 days, with the reductions in FeNO restricted to participants with the highest FeNO at baseline in the urban environment.^31^

Several additional results in our study merit further discussion. We found evidence for larger effects of long-term PM_2.5_ and NO_2_ in females as compared to males (p=0.007 and 0.07, respectively), but no evidence for effect modification by asthma or allergy. In the preliminary CHS analysis of long-term air pollution effects on FeNO, we had found no evidence for effect modification by sex.^16^ Sex/gender differences have been observed previously in air pollution epidemiology analyses, generally with larger effects for boys in early life and girls/women in later childhood and older age.^32^ We also found a statistically significant negative association for long-term O_3_, which was not robust to our various sensitivity analyses. Within-community annual average O_3_ was negatively correlated with PM_2.5,_ PM_10_, and NO_2_ and unlike those pollutants, there were trends of increasing O_3_ during the study period. Finally, the association of long-term NO_2_ with FeNO was larger in magnitude when leaving out one community (Upland) or including a community-level random slope. Upland had the highest annual average NO_2_ at the beginning of the study period, and one of the largest declines over the study period.This study has several strengths, including: a large ethnically diverse population-based cohort of schoolchildren; prospective longitudinal assessment of FeNO (up to 6 measurements over an eight year period); a Southern California study location with substantial within- and between-community variation in ambient air pollution exposures during the study period, representative of the range observed nationally in the United States; as well as a study design and statistical analyses approach that allowed us to target within-person associations of long-term air pollution and FeNO (automatically controlling for time-constant confounders) while adjusting for short-term air pollution exposures.

This study has several limitations. The method for FeNO assessment changed during the study period, as technology improved (offline assessment used for the first two FeNO visits and online assessment used for the next four FeNO visits). Although the offline values were converted to predicted online values based on a published internally-developed prediction model with an *R*^*2*^ = 0.94 ^33^, there remains a possibility that this change in breath collection technique may have impacted our effect estimates. In a sensitivity analysis restricting to only online FeNO visits, the association of long-term NO_2_ with FeNO remained similar and statistically significant (though with wider confidence intervals because of smaller sample size) and the association of long-term PM_2.5_ with FeNO was still positive, but attenuated and no longer statistically significant. A limitation of our study design is that FeNO was assessed in only the most recent CHS cohort, allowing only for investigation of associations with within-community fluctuations in annual average air pollution. We were unable to investigate associations of within-community fluctuations in multi-year averages. Previous high impact CHS publications have taken advantage of multiple CHS cohorts (with staggered recruitment over time from the same communities) to demonstrate that, in the same communities, declines in long-term (multi-year) average air pollution have been associated with improved lung function growth,^34^ reduced bronchitic symptoms,^35^ and reduced asthma incidence.^36^ However, the most recent CHS cohort was still recruited from multiple cities, with a wide range of annual average ambient air pollution levels at baseline and differing trends over the course of the study period.

In conclusion, our findings provide the strongest evidence to date that long-term PM_2.5_ and NO_2_ exposures affect within-participant FeNO, independent of the effects of short-term exposures, even during a study period with declining pollution levels. Longitudinal FeNO measurements may be useful as an early marker of chronic respiratory effects of long-term air pollution exposures in children.

## Data Availability

The data that support the findings of this study are available on request, which will be reviewed and approved by the University of Southern California Health Campus Institutional Review Board.

## Abbreviations

ATS: American Thoracic Society
BMI: body mass index
CHS: Southern California Children’s Health study
EC: elemental carbon
FeNO: Fractional exhaled nitric oxide
NO: Nitric oxide
NO_2_: Nitrogen dioxide
O_3_: Ozone
PM_2.5_: particulate matter with an aerodynamic diameter of 2.5 or less
PM_10_: particulate matter with an aerodynamic diameter of 10 μm or less
SHS: secondhand tobacco smoke

## Acknowledgements

This work was supported by the Office of the Director of the National Institutes of Health (UG3OD023287 and UG3OD023249); the National Institutes of Environmental Health Sciences (grant # R01ES023262, R01ES027860); the Southern California Environmental Health Sciences Center (grant # P30ES007048) funded by the National Institute of Environmental Health Sciences; the Children’s Environmental Health Center (grant #s P01ES009581, R826708-01 and RD831861-01) funded by the National Institute of Environmental Health Sciences and the Environmental Protection Agency; the Maternal and Developmental Risks from Environmental and Social Stressors (MADRES) Center (grant #s P50ES026086, 83615801-0) funded by the National Institute of Environmental Health Sciences, the National Institute for Minority Health and Health Disparities and the Environmental Protection Agency and the Hastings Foundation. We are indebted to the school principals, teachers, students and parents in each of the study communities for their cooperation and especially to the members of the field team for their efforts.

