## Supplementary material for "Long-term Exposures to Air Pollutants Affect FeNO in Children: A Longitudinal Study": Online Supplement

**Affiliations:**
^1^ Department of Internal Medicine, University of Utah, Salt Lake City, UT, USA;

^2^ Department of Family and Preventive Medicine, University of Utah, Salt Lake City, UT, USA;

^3^ Veteran Affairs Salt Lake City Health Care System, Salt Lake City, UT, USA;

^4^ Department of Preventive Medicine, University of Southern California, Los Angeles, CA, USA;

^5^ Department of Biostatistics, Columbia University, New York, NY, USA;

**Corresponding author contact information:**

Yue Zhang

Division of Epidemiology

Department of Internal Medicine

University of Utah

295 Chipeta Way

Salt Lake City, UT, 84018

**ABSTRACT**

Fractional exhaled nitric oxide (FeNO) is a marker of airway inflammation shown to be responsive to short-term air pollution exposures; however, effects of long-term exposures are uncertain. Using longitudinal assessments of FeNO and air pollutant exposures, we aimed to determine whether FeNO is a marker for chronic effects of air pollution exposures after accounting for short-term exposures effects.

FeNO was assessed up to six times 2004-2012 in 3607 schoolchildren from 12 communities in the Southern California Children’s Health Study. Within-community long-term ambient air pollution exposures (PM_2.5_, PM_10_, NO_2_, O_3_) were represented by differences between community-specific annual averages and the eight-year average spanning the study period. Linear mixed-effect models estimated within-participant associations of annual average air pollution with current FeNO, controlling for previous FeNO, prior seven-day average pollution, potential confounders, and community-level random intercepts. We considered effect modification by sex, ethnicity, asthma, and allergy at baseline.

We found FeNO was positively associated with annual average air pollution, after accounting for short-term exposures. One standard deviation higher annual PM_2.5_ and NO_2_ exposures (PM_2.5:_2.0 μg/m^3^; NO_2_:2.7 ppb) were associated, respectively, with 4.6% (95%CI:2.3%-6.8%) and 6.5% (95%CI:4.1%-8.9%) higher FeNO. These associations were larger among females. We found little evidence supporting association with PM_10_ or O_3_.

Annual average PM_2.5_ and NO_2_ levels were associated with FeNO in schoolchildren, adding new evidence that long-term exposure affects FeNO beyond the well-documented short-term effects. Longitudinal FeNO measurements may be useful as an early marker of chronic respiratory effects of long-term PM_2.5_ and NO_2_ exposures in children.

**Supplementary information**

**Statistical Methods**

The general model form is presented below with subscripts *c*, *i*, and *j* indexing community, participant, and visit, respectively:

$$\log\left( {FeNO}_{cij} \right)=\beta_{0}+\beta_{1}\cdot\left( {AP}_{LT,cij}-{AP}_{LT,c} \right)+\beta_{2}\cdot{AP}_{LT,c}+\beta_{3}\cdot{AP}_{ST,cij}+\beta_{4}\cdot\log\left( {FeNO}_{cij-1} \right)+f\left( {Age}_{cij},{Height}_{cij} \right)+\boldsymbol{\gamma\cdot}\boldsymbol{Z}_{\boldsymbol{ci}}+\boldsymbol{\theta\cdot}\boldsymbol{W}_{\boldsymbol{cij}}+e_{c}+\epsilon_{ci}+\varepsilon_{cij}$$

and where ${AP}_{LT,c}$ denotes the seven-year average air pollutant concentration for each community, $\left( {AP}_{LT,cij}-{AP}_{LT,c} \right)$ denotes long-term exposure (i.e., the annual within-community fluctuation), and ${AP}_{ST,cij}$ denotes short term exposure (i.e., seven-day average); $\boldsymbol{Z}_{\boldsymbol{ci}}$ and $\boldsymbol{W}_{\boldsymbol{cij}}$ denote time-independent and time-dependent confounders, respectively; $f\left( {Age}_{cij},{Height}_{cij} \right)$ denotes the FeNO trajectory as a linear combination of $Age$, $Height$ and their interaction; $e_{c}$ and $\epsilon_{ci}$ represent community-level and participant-level random intercepts. Controlling for log FeNO at the previous visit (log ${FeNO}_{cij-1}$) enables better control of the within-participant correlation and controls for any carry-over effects of air pollution exposure prior to the previous visit. The parameter of primary interest is $\beta_{1}$, which quantifies the association between long-term exposure (annual within-community fluctuations) and log FeNO, adjusting for short-term exposure, confounders, and log FeNO at the previous visit.

| **Supplementary Table 1: Adjusted**^*^ **percent difference in FeNO associated with one standard deviation (SD) increases in long-term air pollution exposures**^†^ **in various sensitivity analyses.** | | | | |
| --- | --- | --- | --- | --- |
|  | **Percent difference in FeNO per SD**^‡^ **increase (95% CI) in** | | | |
| **Model** | **PM_2.5_** | **PM_10_** | **NO_2_** | **O_3_** |
| Primary analysis (Table 4) | 4.55 (2.33, 6.82) | 0.63 (-1.88, 3.21) | 6.46 (4.08, 8.90) | -2.62 (-4.49, -0.71) |
| Unadjusted^§^ | 1.95 (0.05, 3.88) | 0.21 (-1.78, 2.23) | 4.04 (1.66, 6.47) | -0.95 (-2.49, 0.62) |
| Excluding 5 (of 12) communities with only 4 (of 6) FeNO visits | 6.04 (3.72, 8.40) | 1.26 (-1.08, 3.66) | 4.78 (2.40, 7.20) | -1.80 (-3.79, 0.23) |
| Including only online FeNO data (last 4 of 6 visits) | 1.82 (-4.27, 8.3) | 4.77 (-1.73, 11.71) | 6.28 (2.22, 10.51) | -4.66 (-12.51, 3.88) |
| Autoregressive error term (lme), no adjustment for previous FeNO | 2.12 (-0.01, 4.29) | 0.21 (-2.18, 2.66) | 2.19 (-0.07, 4.50) | -1.62 (-3.37, 0.16) |
| Adjust for first FeNO (rather than previous visit FeNO) | 1.63 (-0.3, 3.59) | 0.34 (-1.83, 2.56) | 2.50 (0.46, 4.58) | -1.12 (-2.81, 0.59) |
| No adjustment for 7-day average of the same pollutant | 3.28 (0.58, 2.68) | -0.03 (-2.38, 2.37) | 6.37 (4.15, 8.64) | -1.29 (-3.01, 0.46) |
| ^*^Unless otherwise specified, all models adjusted for sex, race, respiratory allergy, asthma, medication use, wheeze, secondhand tobacco smoking, recent respiratory illness, room air nitric oxide, month, temperature, prior 7 day average of the same pollutant, and natural log-transformed FeNO at the previous visit.  ^§^: Only adjusted for basic variables (including sex, race, age, height and natural log-transformed FeNO at the previous visit.  ^†^Long-term exposures are annual within-community fluctuations, calculated as the difference between annual air pollutant concentration and seven-year average concentration.  ^‡^ SDs are: 2.0 μg/m^3^ for PM_2.5_, 6.7 μg/m^3^ for PM_10_, 2.7 ppb for NO_2_ and 2.5 ppb for O_3_.^§^ Presented estimates are for the “average” community, with a random slope of 0. Estimates of the random slope SD are: 0.027 for PM_2.5_, 0.062 for PM_10_, 0.078 for NO_2_ and 0.069 for O_3_.  ^: p-value<0.1 | | | | |

| **Supplementary Table 2: Adjusted**^*^ **percent difference in FeNO associated with a one standard deviation (SD)**^‡^ **increase in long-term air pollution exposures,**^†^ **in sensitivity analysis removing one community at a time.** | | | | |
| --- | --- | --- | --- | --- |
|  | **Percent difference in FeNO (95% CI)** | | | |
| **Removed Community** | **PM_2.5_** | **PM_10_** | **NO_2_** | **O_3_** |
| **1:** Alpine | 5.19 ( 3.05 , 7.37 ) | 1.35 ( -0.83 , 3.59 ) | 5.25 ( 2.89 , 7.68 ) | -1.76 ( -3.64 , 0.16 ) |
| **2:** Lake Elsinore | 4.32 ( 2.08 , 6.61 ) | 0.7 ( -1.84 , 3.30 ) | 6.04 ( 3.64 , 8.51 ) | -2.36 ( -4.26 , -0.43 ) |
| **3:** Lake Gregory | 4.55 ( 2.33 , 6.82 ) | 0.63 ( -1.88 , 3.21 ) | 6.46 ( 4.08 , 8.90 ) | -3.69 ( -5.69 , -1.65 ) |
| **6:** Long Beach | 4.35 ( 2.13 , 6.63 ) | 1.28 ( -1.28 , 3.91 ) | 5.61 ( 3.15 , 8.13 ) | -2.99 ( -4.87 , -1.06 ) |
| **7:** Mira Loma | 6.21 ( 3.52 , 8.97 ) | 0.37 ( -2.30 , 3.12 ) | 6.45 ( 4.00 , 8.95 ) | -1.81 ( -3.86 , 0.28 ) |
| **8:** Riverside | 2.88 ( 0.60 , 5.22 ) | -1.75 ( -4.83 , 1.43 ) | 5.36 ( 2.91 , 7.87 ) | -2.06 ( -4.12 , 0.03 ) |
| **9:** San Dimas | 3.01 ( 0.67 , 5.42 ) | 0.83 ( -1.77 , 3.49 ) | 6.95 ( 4.43 , 9.54 ) | -3.37 ( -5.42 , -1.28 ) |
| **11:** Santa Maria | 4.83 ( 2.59 , 7.12 ) | 0.70 ( -1.83 , 3.30 ) | 6.43 ( 4.07 , 8.84 ) | -2.53 ( -4.41 , -0.62 ) |
| **12:** Upland | 5.93 ( 3.58 , 8.33 ) | 1.62 ( -1.21 , 4.53 ) | 16.36 ( 12.81 , 20.03 ) | -2.94 ( -4.94 , -0.90 ) |
| **13:** Glendora | 3.99 ( 1.52 , 6.53 ) | 0.46 ( -2.22 , 3.21 ) | 5.32 ( 2.64 , 8.07 ) | -1.41 ( -3.58 , 0.81 ) |
| **14:** Anaheim | 4.71 ( 2.44 , 7.04 ) | 2.27 ( -0.41 , 5.02 ) | 7.73 ( 5.23 , 10.29 ) | -2.73 ( -4.35 , -0.36 ) |
| **15:** San Bernardino | 5.22 ( 2.91 , 7.58 ) | 0.90 ( -1.67 , 3.54 ) | 6.55 ( 4.14 , 9.01 ) | -2.56 ( -4.45 , -0.64 ) |
| **16:** Santa Barbara | 4.71 ( 2.31 , 7.16 ) | -0.36 ( -3.01 , 2.36 ) | 5.75 ( 3.25 , 8.32 ) | -3.33 ( -5.22 , -1.39 ) |
| ^*^Models adjusted for: sex, race, respiratory allergy, asthma, medication use, wheeze, secondhand tobacco smoking, recent respiratory illness, room air nitric oxide, month, temperature, prior 7 day average of the same pollutant, and natural log-transformed FeNO at the previous visit.  ^†^Long-term exposures are annual within-community fluctuations, calculated as the difference between annual air pollutant concentration and seven-year average concentration.  ^‡^ SDs are: 2.0 μg/m^3^ for PM_2.5_, 6.7 μg/m^3^ for PM_10_, 2.7 ppb for NO_2_ and 2.5 ppb for O_3_ . | | | | |
